# Serotype specificity and utility of antibodies 5D8/1 and Cox mAB 31A2 for enterovirus diagnostics

**DOI:** 10.1101/2022.08.17.22278922

**Authors:** Jutta E. Laiho, Marie Zeissler, Noel G. Morgan, Heikki Hyöty, Sarah J. Richardson

## Abstract

A commercially available antibody, Cox mAB 31A2, raised against the VP1 protein of coxsackievirus B3 (CVB3) has been reported as suitable for the detection of CVB3 in diagnostic samples (Ettischer-Schmid 2016). The authors compared this antibody with the widely used, multi-specific, monoclonal anti-VP1 antibody marketed by Dako (clone 5D8/1) and concluded that clone 5D8/1 should not be used to identify enterovirus infections in diagnostic samples. Rather they suggested that Cox mAB 31A2 is preferable for this purpose. Here we address these issues and show that Cox mAB 31A2 can be used successfully to diagnose CVB3 infection in various cell and tissue samples but we demonstrate that it fails to detect many clinically relevant enterovirus types, thereby limiting its use as a general diagnostic reagent for clinical specimens. Rather, we propose that, when used under carefully controlled conditions, clone 5D8/1 should remain the reagent of choice for such purposes.

## Introduction

Human enteroviruses comprise over 110 different types, including coxsackie B viruses (CVBs^1^). Most enterovirus infections are mild, but they can also cause severe acute illness and have been linked to more chronic diseases such as various cardiomyopathies and type 1 diabetes. Multiple enterovirus types may be involved in causing these conditions and it is important that screening methods are developed which have an appropriately broad specificity.

Immunohistochemical analysis of tissue samples is frequently employed to detect enterovirus proteins and it has become increasingly popular to employ a broad-spectrum monoclonal antibody (clone 5D8/1, Dako/Agilent) which targets an immunodominant epitope of the CVB5 VP1 structural protein. This antibody is among the most versatile of enteroviral antibodies in the diagnostic arsenal since it recognizes multiple enterovirus species. However, clone 5D8/1 also cross-reacts with a defined subset of cellular proteins under certain conditions [1, 2] which may limit its utility as a diagnostic tool because of the risk of false positive immunostaining. In an attempt to circumvent this problem, a new mouse monoclonal anti-CVB3 VP1 antibody (Cox mAB 31A2, Mediagnost, Germany) has been developed for multiple molecular applications. This reagent is reportedly devoid of non-specific cross-reactivity in cells and tissue samples and has been proposed as a replacement for clone 5D8/1 for use in immunohistochemical analysis of CVB infected cells and tissues. To assess the validity of this conclusion, we sought to compare the specificity and selectivity of Cox mAB 31A2 with clone 5D8/1 under standardized conditions in two independent laboratories.

## Material and Methods

The specificity and sensitivity of Cox mAB 31A2 and clone 5D8/1 were examined using three different formalin-fixed paraffin-embedded (FFPE^2^) cell microarrays (CMAs^3^), two of which (enterovirus CMA and CVB1 CMA) have been described previously [3] while the third was designed to display 9 different CVB3 strains (Supplementary Table S1.). In addition, the relative sensitivity of each antiserum to detect CVB1 infection in cultured A549 cells serially diluted up to 10^−8^ fold was also tested [4, 5].

Human FFPE autopsy tissue from either culture-proven CVB2-or CVB4-infected, or normal neonatal hearts, was used with permission from the West of Scotland Research Ethics Committee. The histological features of these tissues have been described in detail previously [5, 6]. Coxsackie-infected neonatal mice [7] were examined to test the antibody specificity in mouse tissues.

Chromogenic immunohistochemical assays were performed in two independent laboratories, manually in Exeter and with an automated system in Tampere [4, 8]. For Cox mAB 31A2 immunostaining, a final concentration of 1:1000 was employed in both laboratories. Clone 5D8/1 was used at a concentration of 240ng/ml in Tampere and at 55ng/ml in Exeter. Where relevant, a mouse IgG isotype control was applied. Slides were visualized with an Olympus BX60 microscope (Tampere) or a Nikon 50i Microscope fitted with a DS-Fi camera and a DSL2 camera control unit (Exeter).

## Results

Immunohistochemical staining of the CMA infected acutely with enteroviruses revealed that Cox mAB 31A2 readily detects CVB3 infection and also, to a lesser extent, CVB1. Weak recognition of both CVB2 and CVB5 was also observed (Table 1 and Fig 1. A). All other enterovirus types and the uninfected control cell lines were immunonegative. A comparison of the staining patterns achieved with Cox mAB 31A2 and clone 5D8/1, is shown in Table 1 and Figure 1.

**Table 1.**
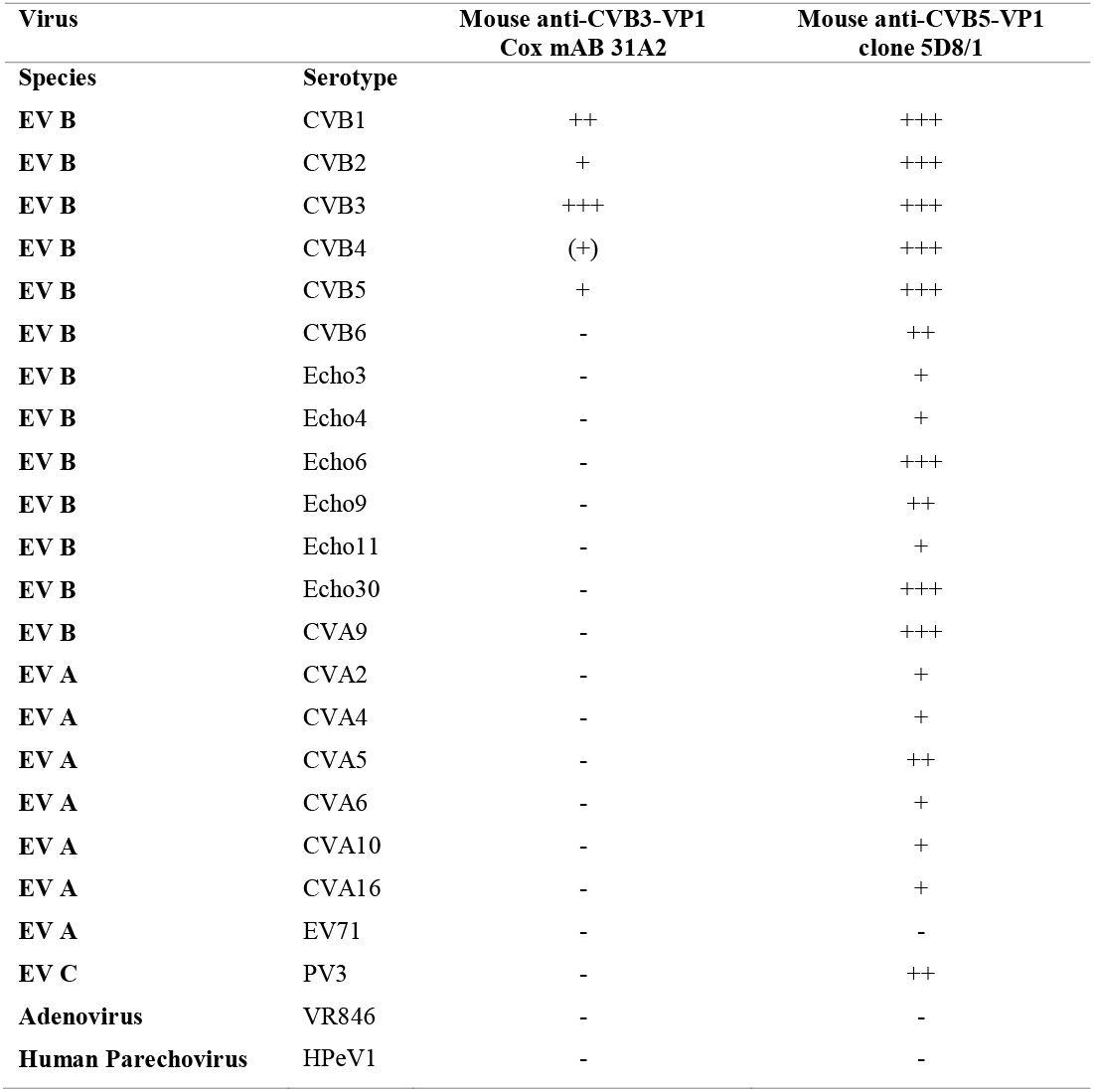
Cox mAB 31A2 recognizes distinct CVB serotypes, while clone 5D8/1 demonstrates a more broad-spectrum specificity, detecting enteroviruses (EVs) from species A, B and C (Previously published in [10]). Recognition scale: +++ = strong, ++ = moderate, + = weak, -= negative.

**Figure 1.**
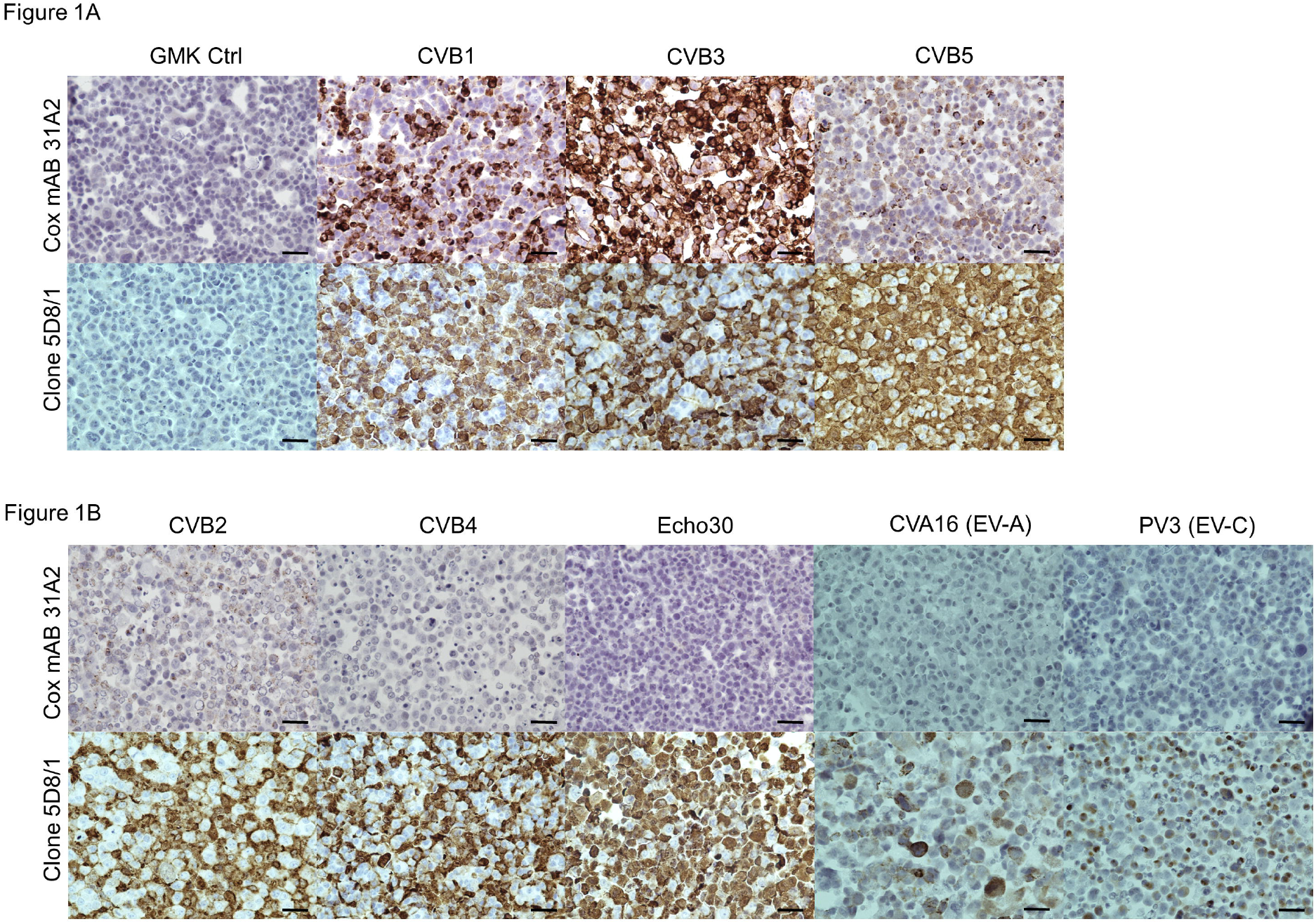
**A**. Cox mAB 31A2 and clone 5D8/1 recognize CVB1, CVB3 and CVB5. Representative images of 31A2 (upper panel) and Clone 5D8/1 (lower panel) staining in acutely-infected CVB1, CVB3, CVB5 and control GMK cells. **B**. Representative images of enteroviruses that are differentially recognized by Cox mAB 31A2 (upper panel) and clone 5D8/1 (lower panel). Scale bar 5μm.

Staining of CVB3 and CVB1 CMAs with Cox mAB 31A2 confirmed that this antibody recognized all (9) of the tested CVB3 wild type strains and all (31) CVB1 strains. In a limiting dilution series with CVB1, a dilution of 10^−3^ was reached before loss of signal with Cox mAB 31A2, while clone 5D8/1 was still effective at a dilution of 10^−6^ [4]. Immunostaining of tissues from neonatal mice infected with CVB1, CVB3 and CVB5 [7] confirmed the acute CMA results, since the order of preference for Cox mAB 31A2 was CVB3>CVB1>CVB5 (data not shown).

Neonatal heart tissue from people with confirmed CVB-infection (CVB2 and CVB4;[5, 6] or those without infection were each treated with Cox mAB 31A2 and clone 5D8/1. No staining was observed in the uninfected heart or in heart with CVB4 infection. A few very faintly positive cells were observed in CVB2-infected tissue. By contrast, clone 5D8/1 yielded a strongly positive signal in CVB2 and CVB4-infected, whilst no staining was observed in the uninfected control heart (Fig 2A-D). To reveal the localization of virus in these tissues, an antiserum directed against double stranded RNA (dsRNA^4^; formed during the replication cycle) was employed in adjacent tissue sections. The labelling pattern achieved with the dsRNA (J2) antiserum [9], was remarkably similar to that obtained with clone 5D8/1 (Fig 2F). No staining was observed in infected heart tissue when the J2 antibody was pre-incubated with polyIC (a dsRNA mimic; Fig 2G).

**Figure 2.**
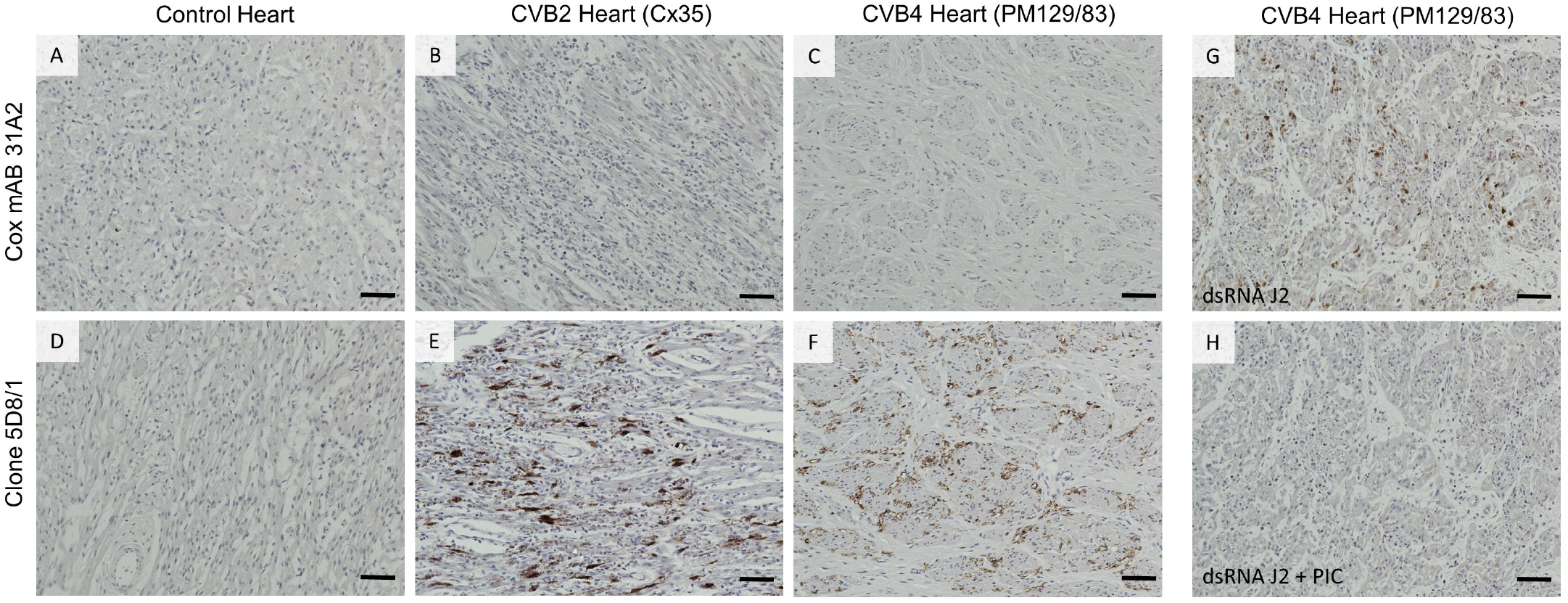
Representative images of Cox mAB 31A2 (upper panel A-C) and clone 5D8/1 (lower panel D-E) staining in control heart (A&D) and heart from neonates with a lethal culture confirmed CVB infection, CVB2 (B&E) or CVB4 (C&F). Representative images of dsRNA staining in CVB4-infected heart (G) confirms the presence of virus in this case. Pre-incubation of the dsRNA J2 antisera with polyIC (PIC; a dsRNA mimic) confirms the specificity of the staining as described previously [9]. Scale bar 40μm.

## Discussion

Within Pathology, there remains an urgent need to develop an arsenal of broad spectrum reagents which are able to verify a suspected diagnosis of enteroviral infection and such reagents should be capable of detecting the majority of enteroviral serotypes with high specificity and sensitivity. Accordingly, one antibody in particular, Dako clone 5D8/1, has become increasingly popular for use in screening studies. This reagent was raised against heat-inactivated CVB5, and its epitope is a short peptide within the capsid protein VP1 [10, 11]. Importantly, it also detects the VP1 proteins produced by a broad range of additional enterovirus serotypes but, in addition, clone 5D8/1 may cross-react with certain cellular proteins [2]. Potentially, such cross-reactivity could compromise its utility as a general screening reagent and, as a consequence, continuing attempts are in hand to develop additional antibodies with improved specificity and selectivity. One such is the monoclonal antibody Cox mAB 31A2 which was raised against an epitope present within the VP1 capsid protein of CVB3 and was shown to detect this protein with minimal evidence of cross-reactivity against cellular antigens [1].

In the present study, we have employed Cox mAB 31A2, alongside clone 5D8/1, to compare their accuracy and sensitivity in detecting enteroviral infection in cultured cells, mouse and human tissue samples. The results demonstrate that Cox mAB 31A2 is efficient and selective as a detection reagent when specific enteroviral serotypes (CVB3 and, to a lesser extent, CVB1) are present in the infected material but they also imply that such a narrow specificity might preclude its use for broad-spectrum screening purposes. Set against this, we find that clone 5D8/1 can be used without significant evidence of non-specific immunoreactivity [11] and that, unlike Cox mAB 31A2, it detects a wide range of enterovirus serotypes in cultured cell samples and in heart tissue recovered from individuals with culture-proven CVB infection. On this basis, it would be premature to consider the wholesale replacement of 5D8/1 with Cox mAB 31A2 for broad spectrum screening purposes or to summarily relegate clone 5D8/1 from the arsenal of reagents used to monitor enterovirus infection, where the serotype is unknown.

As noted above, the principal issue that arises most often in relation to the use of clone 5D8/1 is its potential cross-reactivity with native proteins present in tissue or cell samples. Such cross-reactivity has been well documented; including by ourselves. Indeed, we confirm that, in our hands, non-specific immunostaining can be observed quite readily in both human heart and pancreas under poorly optimized conditions [11]. However, when employed under optimal conditions, neither uninfected cultured cell lines nor control samples of human heart were immunopositive following exposure to clone 5D8/1. In our studies, this was equally true when Cox mAB 31A2 was used. Therefore, these results imply that neither 5D8/1 nor Cox mAB 31A2 displays strong cross-reactivity with endogenous cellular proteins under the conditions we have employed and that each has the potential to be useful as a screening reagent.

Importantly, when assessing the sensitivity of the two anti-VP1 reagents in parallel, it became clear that clone 5D8/1 detected the presence of enteroviral capsid proteins in cultured cells more effectively than Cox mAB 31A2 when serotypes were used which varied from those employed to generate each antiserum. Thus, when probing cells infected with CVB1 (which was not the serotype used to raise either Cox mAB 31A2 or 5D8/1) in a sequential limiting dilution series, selective immunopositivity was retained at much higher dilutions with 5D8/1.

Taken together, the present data confirm that the reagent, Cox mAB 31A2, may prove useful for detection of CVB3 (and, to a lesser extent, CVB1) in infected cell and human tissue samples. However, the data also imply that clone 5D8/1 still offers the greater utility as a screening reagent under conditions where the enteroviral serotype is unknown. This is because it recognizes the VP1 proteins produced by a wider spectrum of enteroviral serotypes and is able to label these with high specificity and selectivity, when used appropriately.

## Data Availability

All data produced in the present study are available upon reasonable request to the authors

## Acknowledgements

We thank Eini Eskola, Eeva Tolvanen and Anne Karjalainen from Faculty of Medicine and Life Sciences, University of Tampere, for technical assistance.

## Footnotes

1 CVB, Coxsackie B virus

2 FFPE, formalin-fixed paraffin-embedded

3 CMA, cell microarray

4 dsRNA, double stranded RNA

## References

1. Ettischer-Schmid N, Normann A, Sauter M, Kraft L, Kalbacher H, Kandolf R, Flehmig B, Klingel K (2016) A new monoclonal antibody (Cox mAB 31A2) detects VP1 protein of coxsackievirus B3 with high sensitivity and specificity. Virchows Arch 469:553–562

2. Hansson SF, Korsgren S, Ponten F, Korsgren O (2013) Enteroviruses and the pathogenesis of type 1 diabetes revisited: cross-reactivity of enterovirus capsid protein (VP1) antibodies with human mitochondrial proteins. J Pathol 229:719–728

3. Laiho JE, Oikarinen S, Oikarinen M, Larsson PG, Stone VM, Hober D, Oberste S, Flodström-Tullberg M, Isola J, Hyöty H (2015) Application of bioinformatics in probe design enables detection of enteroviruses on different taxonomic levels by advanced in situ hybridization technology. J Clin Virol 69:165–171

4. Laiho JE, Oikarinen M, Richardson SJ, Frisk G, Nyalwidhe J, Burch TC, Morris MA, Oikarinen S, Pugliese A, Dotta F, Campbell-Thompson M, Nadler J, Morgan NG, Hyöty H (2016) Relative sensitivity of immunohistochemistry, multiple reaction monitoring mass spectrometry, in situ hybridization and PCR to detect Coxsackievirus B1 in A549 cells. J Clin Virol 77:21–28

5. Hilton DA, Variend S, Pringle JH (1993) Demonstration of Coxsackie virus RNA in formalin-fixed tissue sections from childhood myocarditis cases by in situ hybridization and the polymerase chain reaction. J Pathol 170:45–51

6. Richardson SJ, Willcox A, Bone AJ, Foulis AK, Morgan NG (2009) The prevalence of enteroviral capsid protein vp1 immunostaining in pancreatic islets in human type 1 diabetes. Diabetologia 52:1143–1151

7. Hilton DA, Day C, Pringle JH, Fletcher A, Chambers S (1992) Demonstration of the distribution of coxsackie virus RNA in neonatal mice by non-isotopic in situ hybridization. J Virol Methods 40:155–162

8. Krogvold L, Edwin B, Buanes T, Frisk G, Skog O, Anagandula M, Korsgren O, Undlien D, Eike MC, Richardson SJ, Leete P, Morgan NG, Oikarinen S, Oikarinen M, Laiho JE, Hyoty H, Ludvigsson J, Hanssen KF, Dahl-Jorgensen K (2015) Detection of a low-grade enteroviral infection in the islets of langerhans of living patients newly diagnosed with type 1 diabetes. Diabetes 64:1682–1687

9. Richardson SJ, Willcox A, Hilton DA, Tauriainen S, Hyoty H, Bone AJ, Foulis AK, Morgan NG (2010) Use of antisera directed against dsRNA to detect viral infections in formalin-fixed paraffin-embedded tissue. J Clin Virol 49:180–185

10. Samuelson A, Forsgren M, Sallberg M (1995) Characterization of the Recognition Site and Diagnostic Potential of an Enterovirus Group-Reactive Monoclonal-Antibody. Clin Diagn Lab Immunol 2:385–386

11. Richardson SJ, Leete P, Dhayal S, Russell MA, Oikarinen M, Laiho JE, Svedin E, Lind K, Rosenling T, Chapman N, Bone AJ, Foulis AK, Frisk G, Flodstrom-Tullberg M, Hober D, Hyoty H, Morgan NG (2014) Evaluation of the fidelity of immunolabelling obtained with clone 5D8/1, a monoclonal antibody directed against the enteroviral capsid protein, VP1, in human pancreas. Diabetologia 57:392–401

