## Supplemental Table S1 for "Serotype specificity and utility of antibodies 5D8/1 and Cox mAB 31A2 for enterovirus diagnostics"

**Table S1. CVB3 cell microarray.** Details of the wild-type CVB3 strains used in the study

| Name on the cell microarray | CVB3 Strain/PicoBank No. | Cell line |
| --- | --- | --- |
| CBV3 ATCC | ATCC | A549 |
| CVB3_1 | PB-CBV3_DIPP1.1/02 | A549 |
| CVB3_2 | PB-CBV3_DIPP1.2/02 | A549 |
| CVB3_3 | PB-CBV3_DIPP3/99 | A549 |
| CVB3_4 | PB-CBV3_DIPP4/02 | A549 |
| CVB3_5 | PB-CBV3_DIPP5/01 | A549 |
| CVB3_6 | PB-CBV3_VDP5/5/02 | A549 |
| CVB3_7 | PB-CBV3_VDP6/LK12/01 | A549 |
| CVB3_8 | PB-CBV3_VDP6/LK7/01 | A549 |
| CVB3_9 | PB-CBV3_V13D4C/10 | A549 |
